# Prevalence and risk factors associated with *Haemophilus ducreyi* cutaneous ulcers in Cameroon

**DOI:** 10.1101/2023.07.28.23293301

**Authors:** Philippe Ndzomo, Serges Tchatchouang, Earnest Njih Tabah, Theophilus Njamnshi, Mireille Victorine Noah Tsanga, Jude Alexis Bondi, Rebecca Handley, Camila González Beiras, Jules Tchatchueng, Claudia Müller, Simone Lueert, Sascha Knauf, Onana Boyomo, Emma Harding-Esch, Oriol Mitja, Tania Crucitti, Michael Marks, Sara Eyangoh

## Abstract

Epidemics of yaws-like cutaneous ulcers are regularly documented in children in the tropics. They occur mainly in poor and remote communities without access to health facilities. The integration of molecular tools into yaws control efforts has made it possible to describe *Haemophilus ducreyi* (*HD*) as a major cause of cutaneous ulcers. The objective of this work was to determine the prevalence of *HD* as cause of cutaneous ulcers, as asymptomatic carriage and the risk factors associated.

A cross-sectional study was conducted in yaws endemic districts of Cameroon. Participants included people presenting yaws-like ulcers and asymptomatic individuals. Swab samples were collected from each participant and tested for *HD* and *Treponema pallidum* (*TP*) using established qPCR method. Additionally, demographic, habitat, proximity, and hygiene characteristics were collected using a structured questionnaire.

A total of 443 individuals, including 271 ulcer cases and 172 asymptomatic contacts, were enrolled in this study. The prevalence of *HD* in ulcers was 30.3% (Confidence Interval (CI) 95% [24.8 – 35.7]) and the prevalence of asymptomatic *HD* carriage was 8.6% (CI95% [4.5 – 12.9]). *TP* was also detected in our sample among ulcer cases but in lower proportion (5.2% CI95% [2.5 – 7.8]) compared to *HD*. The adjusted logistic regression model showed that women were as much at risk of having *HD* cutaneous ulcer as men regardless of age; physical proximity to a confirmed ulcer case was the major factor favouring *HD* transmission. *HD* ulcers were more likely to be present on Bantu individuals compare to Baka as well as *HD* colonization.

Data from this study highlight *HD* as the most common cause of cutaneous ulcers in yaws-endemic communities in Cameroon. The real issues of *HD* detection on intact skin are not yet clear. Further studies are needed to elucidate the implications of this carriage in the spread dynamics of the disease.

**Author summary:** Cutaneous ulcers are commonly found affecting children in low-income countries of Africa and the South Pacific. In rural and remote communities of Cameroon the limited access to health care and shortage of sanitation is associated with a high morbidity of cutaneous ulcers. The latter represent an important cause of suffering and stigmatization in children. *HD* is commonly known as the causative agent of the sexually transmitted infection chancroid and has been recently described as a leading cause of cutaneous ulcers in yaws-endemic regions. In this study, we investigated the presence of *HD* and the associated risk factors. Our findings indicate a prevalence of *HD* associated with cutaneous ulcers of 30.3% and a prevalence of asymptomatic *HD* carriage of 5.2%. Physical proximity to a confirmed ulcer case, Bantu ethnicity, and the use of traditional latrines were the main risk factors associated with *HD* ulcers. *TP* DNA was detected in some cutaneous ulcer samples but in lower proportion compared to *HD*. This study confirms that *HD* is a leading cause of cutaneous ulcer in yaws endemic areas in Cameroon. National control programmes in endemic countries should therefore consider this pathogen in their strategies for controlling and eliminating skin neglected tropical diseases (NTDs).

## Introduction

Cutaneous ulcers remain a public health problem in many parts of the world especially in the South Pacific, South East Asia, and West and Central Africa [1–3]. They predominantly affect children living in remote communities with limited water and hygiene and who lack access to health services. The clinical manifestations of cutaneous ulcers vary depending on the causative organism and can range from small sores to multiple deep lesions which can evolve to bone damage [1].

In Africa, South East Asia and the South Pacific Islands most cutaneous ulcers in children have been attributed to yaws, caused by *Treponema pallidum* subsp*. pertenue* (*TPE*) [4,5] based on a clinical diagnosis which is known to be unreliable [6–8]. PCR which is the gold standard diagnosis has made it possible to detect other aetiological agents of cutaneous ulcers such as *HD*, which is often co-endemic and may also occur as a co-morbidity with *TPE*.

*HD* is a fastidious Gram negative coccobacillus traditionally considered as the causative agent of chancroid, a sexually transmitted infection characterized by the appearance of ulcers on the genitals accompanied by suppurative lymphadenopathy [9]. In recent years, multiple studies have reported cutaneous ulcers caused by *H. ducreyi*, including Papua New Guinea [1], [10], the Solomon Islands [11], the Fidji Islands [12], Vanuatu [13], Indonesia [14], Sudan [15], Ghana [16] and Cameroon [17].

The true extent of the burden of *HD* cutaneous ulcers is still unclear, but several studies have established that *HD* may account for 20-60% of skin ulcers clinically diagnosed as yaws [1] [10] [18–20]. Moreover, *HD* was found colonizing fomites and the healthy skin of asymptomatic contacts [21].

The epidemiology of *HD* remains poorly understood. In most endemic areas of Africa and the South Pacific, the prevalence of *HD* as a cause of genital or cutaneous ulcers is not known due to the lack of confirmatory diagnosis in most settings. Traditionally diagnosis relied on culture but this was challenging due to special conditions required for culture which were not available in low and middle income country health services [22]. Today, nucleic acid amplification tests (NAATs) represent an alternative approach. Whilst they still require infrastructure and well-trained personnel, the stringent sample transport requirements which create a barrier to culture *HD* are not needed for NAATs [23,24].

Several factors have been associated with the transmission of certain skin NTDs such as yaws; these include demographic characteristics (age, sex), hygiene and sanitation indicators (poor hand-washing habits, sharing of clothes, frequency of bathing and contact with infected individuals) and housing characteristics (inadequate sanitation, overcrowded houses) [25], but we lack information on the risk factors for *HD* cutaneous ulcers. The limited information that is available suggests there may be both an environmental reservoir and a role for vectors in transmission and these may partly explain the persistence of *HD* ulcers following azithromycin mass drug administration [21].

To address these gaps, we designed a study to determine the prevalence of *HD* as a cause of cutaneous ulcers and the frequency of asymptomatic carriage in yaws endemic districts of Cameroon and to identify associated risk factors. These districts were mainly made up of two major ethnic groups: the Baka and the Bantu. The Baka, also known as pygmies, are a semi-sedentary group of people who live in forest camps and sleep in mud or straw huts. They live essentially from hunting, gathering and, to a lesser extent, from farming, for which they are employed as labourers by their Bantu neighbours, who are sedentary and form a more developed community with modern behaviour [26].

## Methods

### Ethic statement

This study was approved by the National Ethics Committee for Human Health Research (N°2020 / 12/ 1327/ CE/ CNERSH/ SP) and the Ministry of public Health (approval N°631-021). All participants or their legal representatives provided written informed consent.

### Study setting and selection of participants

We conducted a cross-sectional study from May 2021 to May 2022 in 14 health districts in four regions of Cameroon which have been identified as endemic for yaws by the National Yaws, Leishmaniasis, Leprosy and Buruli Ulcer Control Programme (CNLP2LUB). The selected health districts included: Messamena, Abong-Mbang, lomié, Yokadouma, Mbang, Doumé, Batouri, Bétaré-oya, Ndélélé in the East region; Sangmelima, Djoum, Lolodorf in the South region, Bankim in the Adamaoua region and Maroua in the Far North region (Fig 1). Twelve of the 14 health districts participated in both the prevalence and risk factor survey, and the remaining two districts, Maroua and Ndélélé, only participated in the prevalence survey as the study overlapped with the field surveillance activities of the national control programme, which conducted the investigations at these sites.

**Fig 1.**
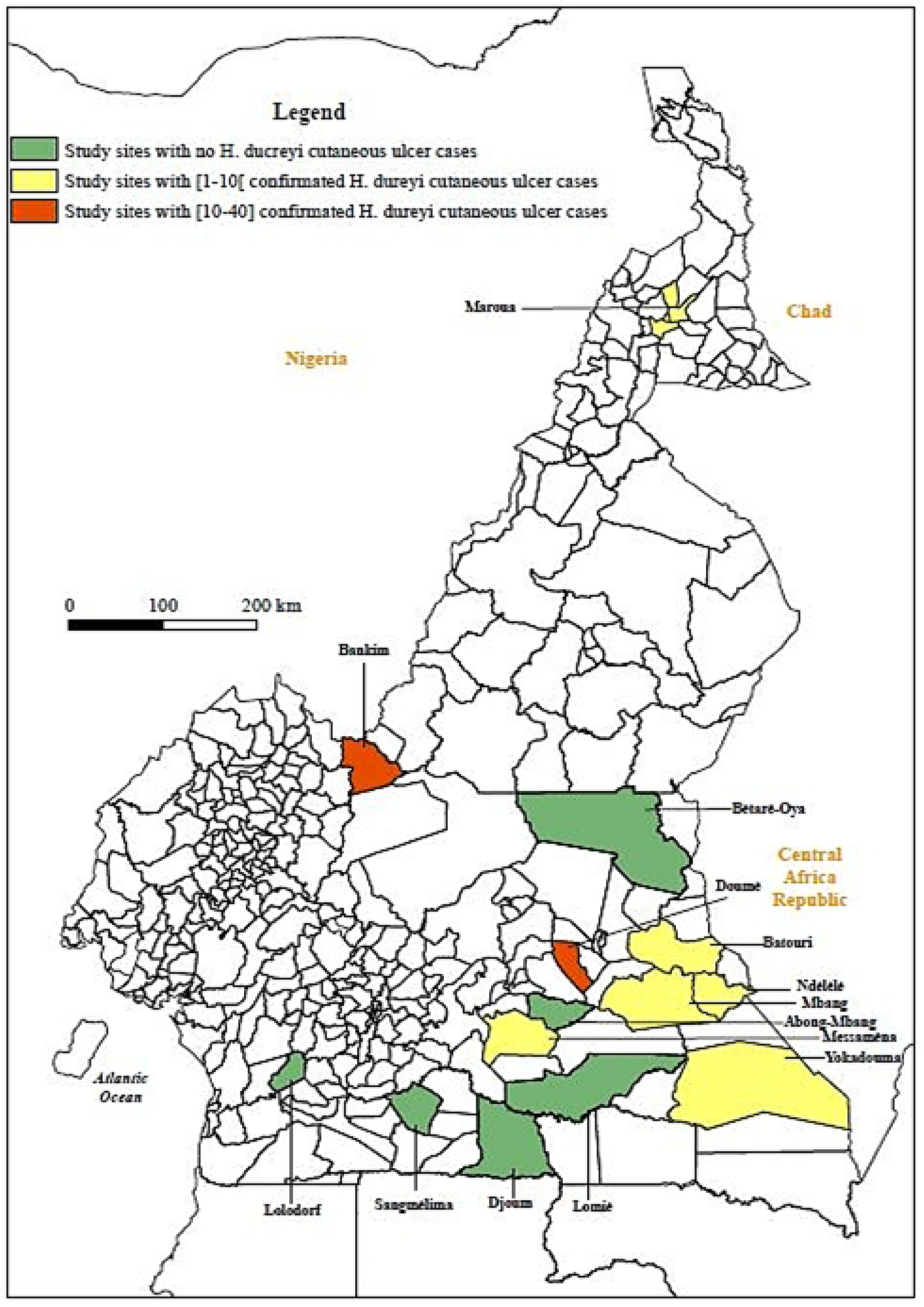
Geographic location of sampling sites and sites with *HD* confirmed ulcer cases.

We carried out an active case searching for cutaneous ulcer cases using both school and community-based activities. For communities with high school enrolment rate and whose screening period coincided with the school year, the case searching was based in schools. For communities with low enrolment rates or where the survey was conducted during the holidays, the case search was house-to-house.

Participants underwent clinical examination and those with cutaneous ulcers were identified and enrolled in the study as cases. Asymptomatic individuals living in the vicinity of the cases, such as the same household or classroom, were enrolled as controls. In addition, we also enrolled asymptomatic participants from households without cases of cutaneous ulcers as a further set of controls.

### Sample collection

We collected samples from cutaneous ulcer cases by swabbing of the ulcerated lesions and from asymptomatic participants by swabbing the front face of both legs with polyester tipped swabs with polystyrene handle (Puritan sterile swabs, Cat #25-806 1PD, Puritan Medical Products-Guilborg, UK). We placed the samples into 500 µL of a custom-made lysis buffer (10 mM Tris, pH 8.0, 0.1 M EDTA, pH 8.0 and 0.5% SDS) and kept them refrigerated (2-8°C) until transport to the Centre Pasteur du Cameroun (CPC) in a cooler, where they were stored at –20°C.

### Laboratory assessment

We extracted genomic DNA from 200 μL of swab lysate using the QIAmp DNA mini kit (Qiagen, Germany) according to the manufacturer’s recommendations and eluted it in a total volume of 100 µL.

We performed on the samples three real time singleplex PCRs on the ABI prism 7500 thermocycler (Thermo Fisher Scientific, Waltham, USA). We first detected the *RNAse P* gene, which codes for an endoribonuclease present in all living cells, and used previously described primers and probe [27] to confirm the adequacy of the sample and the absence of amplifications inhibitors. We targeted the V8 region of the 16S ribosomal RNA gene to detect the presence of *HD* DNA in the samples. The PCR reaction was performed in a total volume of 25 µL containing 0.9 μM of each primer and probe (Hd16SV8-F: 5’>TATACAGAGGGCGGCAAACC<3’; Hd16SV8-R: 5’>CCAATCCGGACTTAGACGTAC<3’; Hd16SV8-P: 5’>Fam-CAAAGG GGAGCGAATCTCAC-BHQ1<3’), ABI TaqMan Fast advanced Master Mix and 2 µL of the DNA template. The cycling conditions were: UNG incubation at 50 °C for 2 minutes, pre-denaturation at 95 °C followed by 40 cycles of denaturation 95 °C for 15 seconds, annealing and extension at 60 °C for 30 seconds. We detected the presence of *TP* by amplifying the *polA* gene (*tp0105*), which is a fragment conserved in all *TP* subspecies, using a protocol described elsewhere [17].

### Questionnaire

We administered a questionnaire to each participant (or their legal representative). We collected data either via a smartphone into ODK collect [28] or via a paper form. The questionnaire consisted of demographic characteristics, information about the house in which the participant lived, data on possible exposure to cases of cutaneous ulcers and measures of access to hygiene and sanitation. Local community health workers acted as a translator where necessary for participants who did not speak English or French.

### Data management and analyses

Each participant was attributed a unique ID code at the moment of inclusion, and that code was used to label his/her risk factor form and samples. The unique ID codes were used for management and analysis of all data related to the participants.

We calculated the prevalence of *HD* as a cause of cutaneous ulcers in the study population and how this varied by key explanatory variables. We then performed three analyses. Firstly, we compared individuals with and without a cutaneous ulcer; secondly, we compared characteristics of individuals whose cutaneous ulcers were or were not caused by *HD*; finally, we compared the characteristics of asymptomatic individuals who were or were not identified as carriers of *HD*. For each comparison we initially fitted a univariate logistics regression model.

We then fitted multivariable models adjusting for age, sex and variables which were significant in the univariate analysis. All analysis were done with SPSS Statistics 20.

## Results

### Description of the population

A total of 24,610 people were clinically screened for cutaneous ulcers at the 14 study sites. Overall, we enrolled 443 participants including 271 individuals with a cutaneous ulcer and 172 asymptomatic contacts (controls). The sex ratio of males to females was 2.1 for ulcer cases and 1.4 for asymptomatic individuals. The age of participants ranged from 2 to 60 years, with a median age of 9 years (Inter Quartile Range (IQR) 7-11).

The overall prevalence of cutaneous ulcers in this population was therefore estimated to be 1.1%. Of these *HD* was identified in 82 (30.3% – 95% CI 24.8 – 36.1%) giving a population prevalence of *HD* ulcers of 0.3%. Eight out of 14 study sites had at least one cutaneous ulcer associated with *HD* (Fig 1). The proportion of ulcers caused by *H. ducreyi* was varied: with Doumé, Maroua and Bankim having the highest proportions of 73.3 % (33/ 45), 72.7 % (8/ 11) and 68.2% (30/ 44) respectively. Amongst asymptomatic controls *HD* was detected in 15 out of 172 participants (8.7% – 95% CI 5 – 14%) (Table 1). All cases of asymptomatic colonisation were detected in only two sites, Bankim (14/15) and Yokadouma (1/15) (S1 Table).

**Table 1:** Proportion of individuals in whom *HD* and *TP* were detected.

| Germ detected | Individuals with a cutaneous ulcer<br>N = 271 | Asymptomatic individuals<br>N = 172 |
| --- | --- | --- |
| *HD* | 82 (30.3 %) | 15 (8.7 %) |
| *TP* | 14 (5.2 %) | 1 (0.6 %) |

Of 271 individuals with a cutaneous ulcer *TP* DNA was detected in 14 (5.2% – 95% CI 2.9 – 8.5%). Cases of yaws were found in only two health districts Mbang (13/14) and Lomié (1/14) and 92.9% were from Baka communities (13/14). Treponemal DNA was detected only in one asymptomatic individual (0.6% – 95% CI 0 – 3.2%) from Mbang (Fig 2).

**Fig 2.**
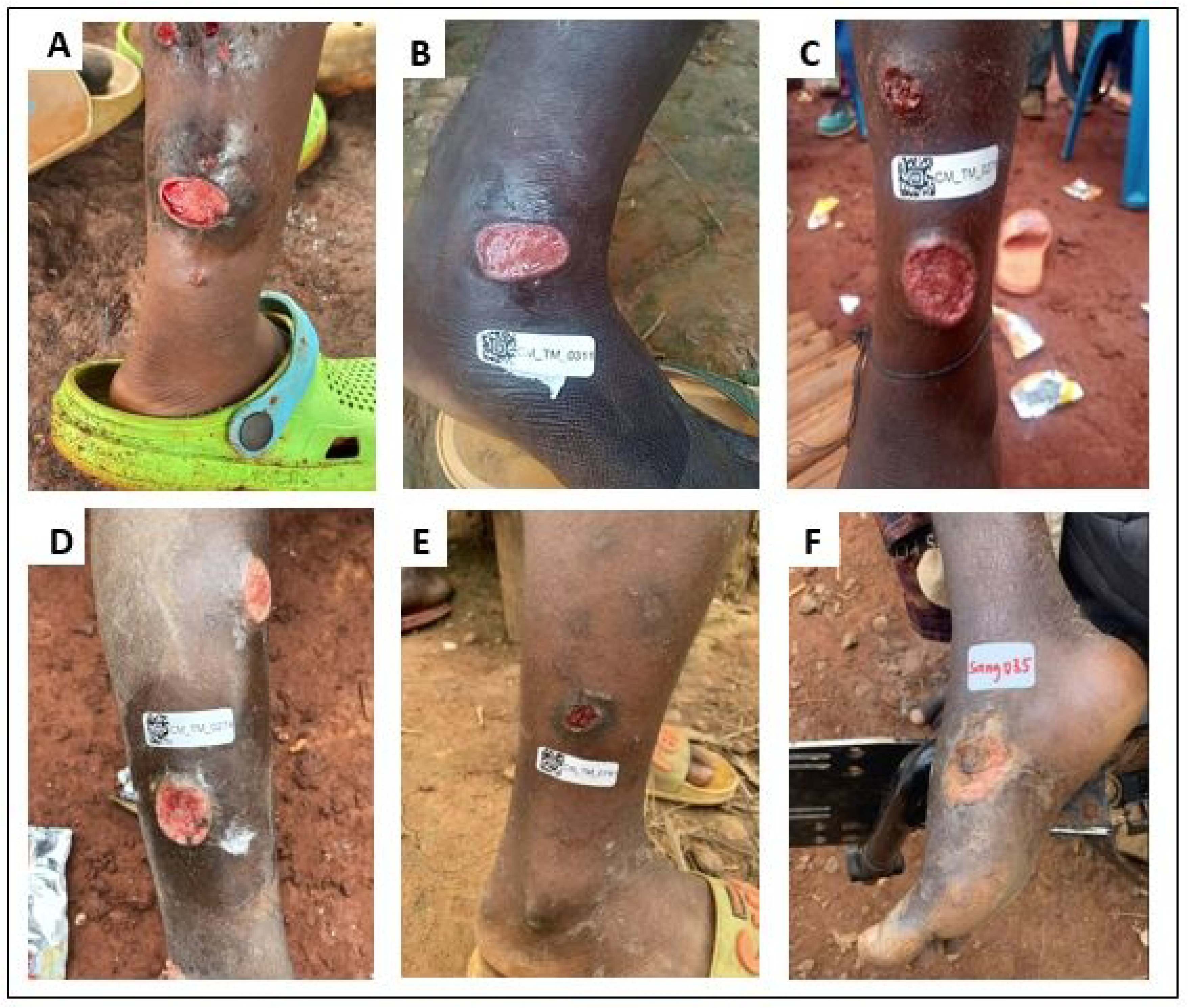
Appearance of cutaneous ulcers similar to yaws in children in Cameroon. Legend: A, B, C, D: *HD* cutaneous ulcers; E: yaws ulcer; F: idiopathic ulcer (negative for both *TP* and *HD*)

Of the 443 participants enrolled in the prevalence study 245 (55.3%) being 127 individuals with ulcers and 118 asymptomatic individuals, completed the risk factor questionnaire and were included in the risk factor analysis.

Both univariate and multivariable analysis revealed that children aged 0-4 years (adjusted Odd Ratio (aOR): 2.91 – 95% CI 1.02– 8.34, p = 0.046) and those aged 15 years and over (aOR: 5.94 – 95% CI 1.15– 30.70, p = 0.034) were more likely to have a cutaneous ulcer (Table 2). Females were about half less likely to have a cutaneous ulcer (aOR: 0.51 – 95% CI 0.28– 0.93, p= 0.028). Members of the Bantou ethnic group had increased odds of yaws like ulcer compared to Baka (aOR = 0.28 – 95% CI 0.14– 0.59, p = 0.001). Some hygiene factors were associated with a reduced risk of cutaneous ulcers including not sharing their clothes with others (aOR: 0.45 – 95% CI 0.25– 0.81, p= 0.008), and living a large distance from a bin (aOR: 0.37 – 95% CI 0.20– 550.69, p= 0.002) but many other household and hygiene related variables did not appear to be associated (Table 2).

**Table 2:**
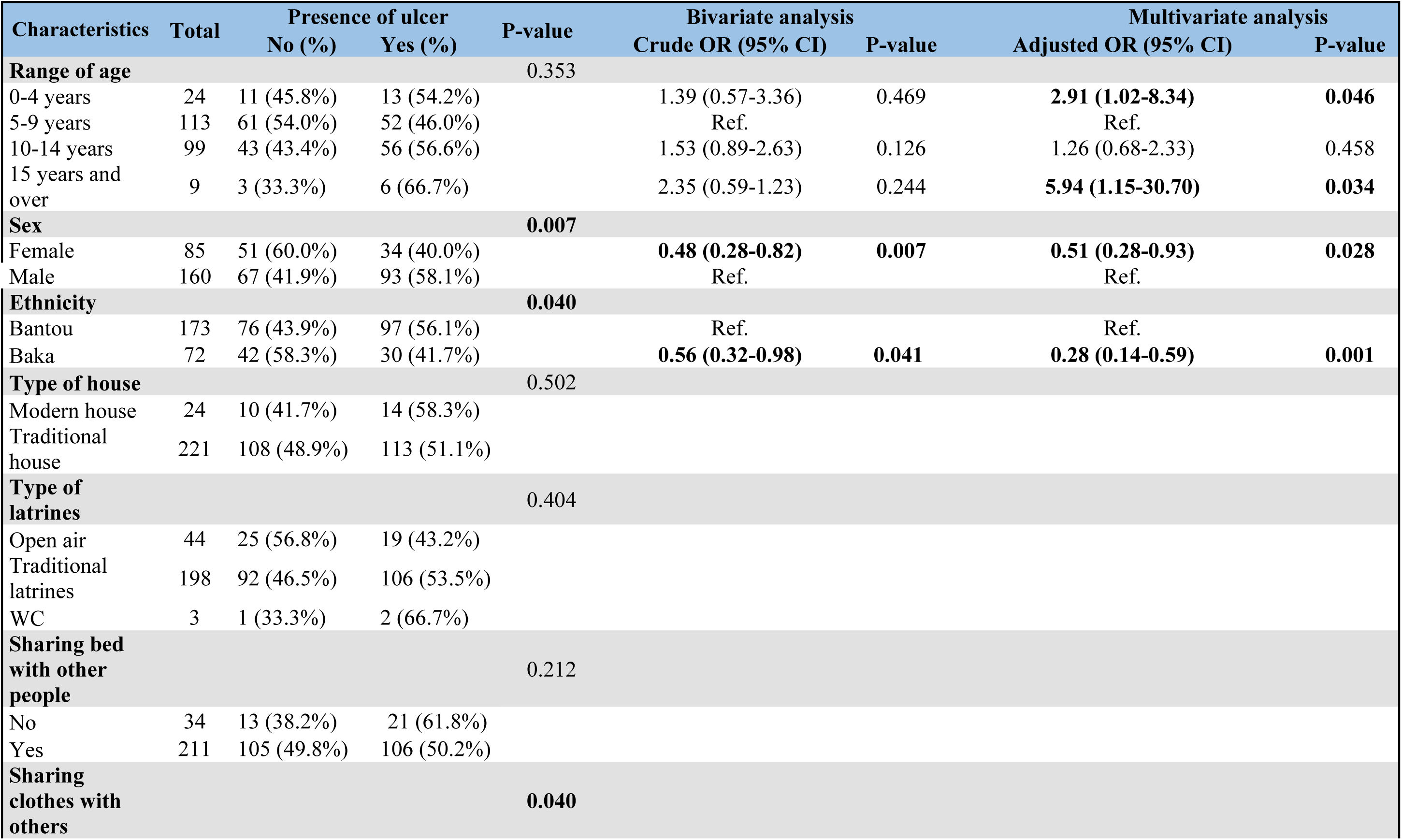
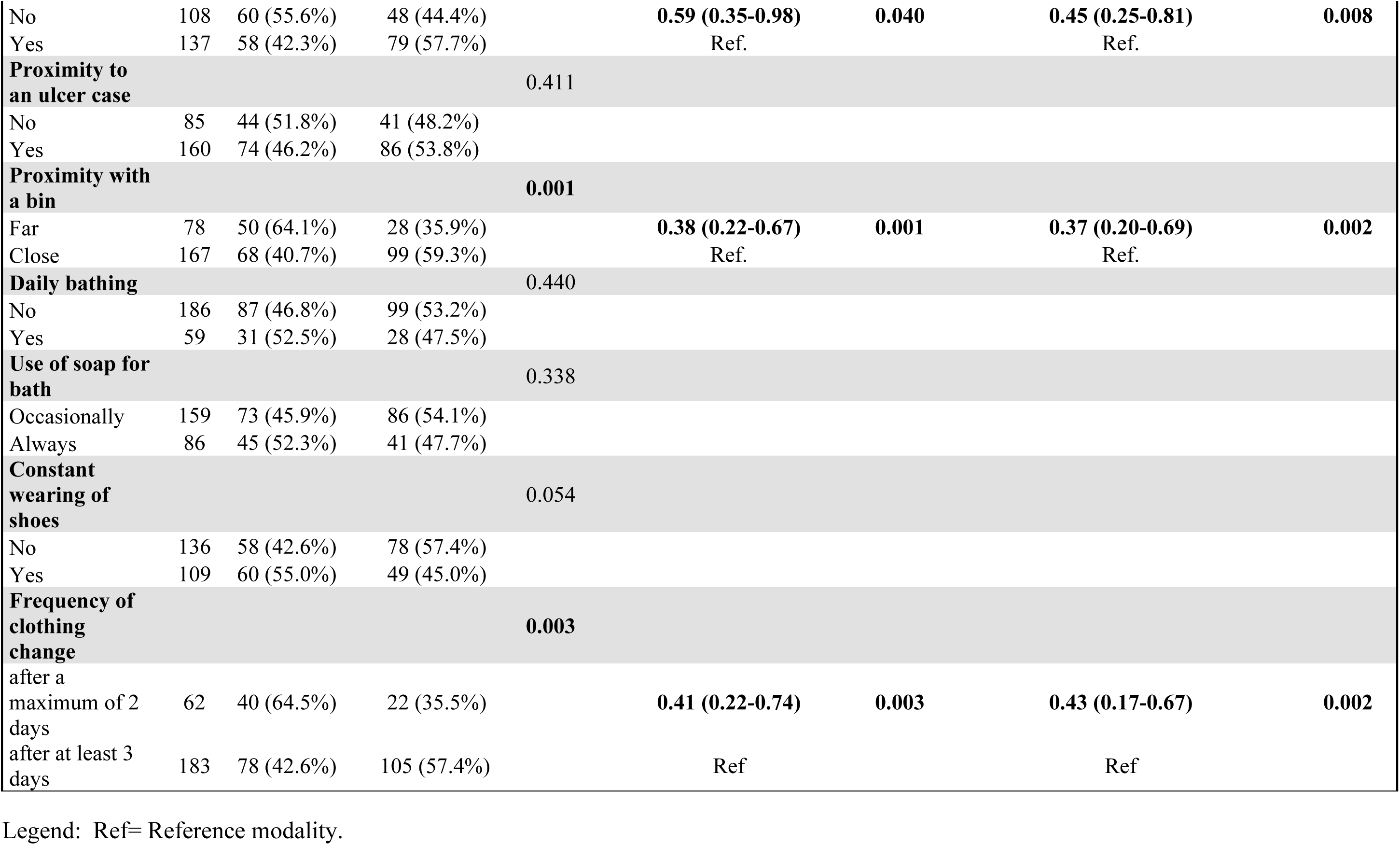
Risk factors associated to yaws like cutaneous ulcers.

Among individuals with cutaneous ulcers, we found that members of the Bantu ethnic group were more likely to have *HD* as the causative agent than Baka (51.5% vs 0%, p= 0.001) as well as people who relieved themselves in traditional latrines (98% (50/51) of confirmed cases used traditional toilets) (Table 3). In addition, there was some evidence that close contact with a cutaneous ulcer case was associated with both *HD* cutaneous ulcer (aOR: 0.27-95% CI 0.11-0.68, p= 0.005) and asymptomatic carriage (close: 20.3% vs not close: 0%, p= 0.005). We also found that the 15 asymptomatic *HD* positive cases were all Bantu (Table 4).

**Table 3.**
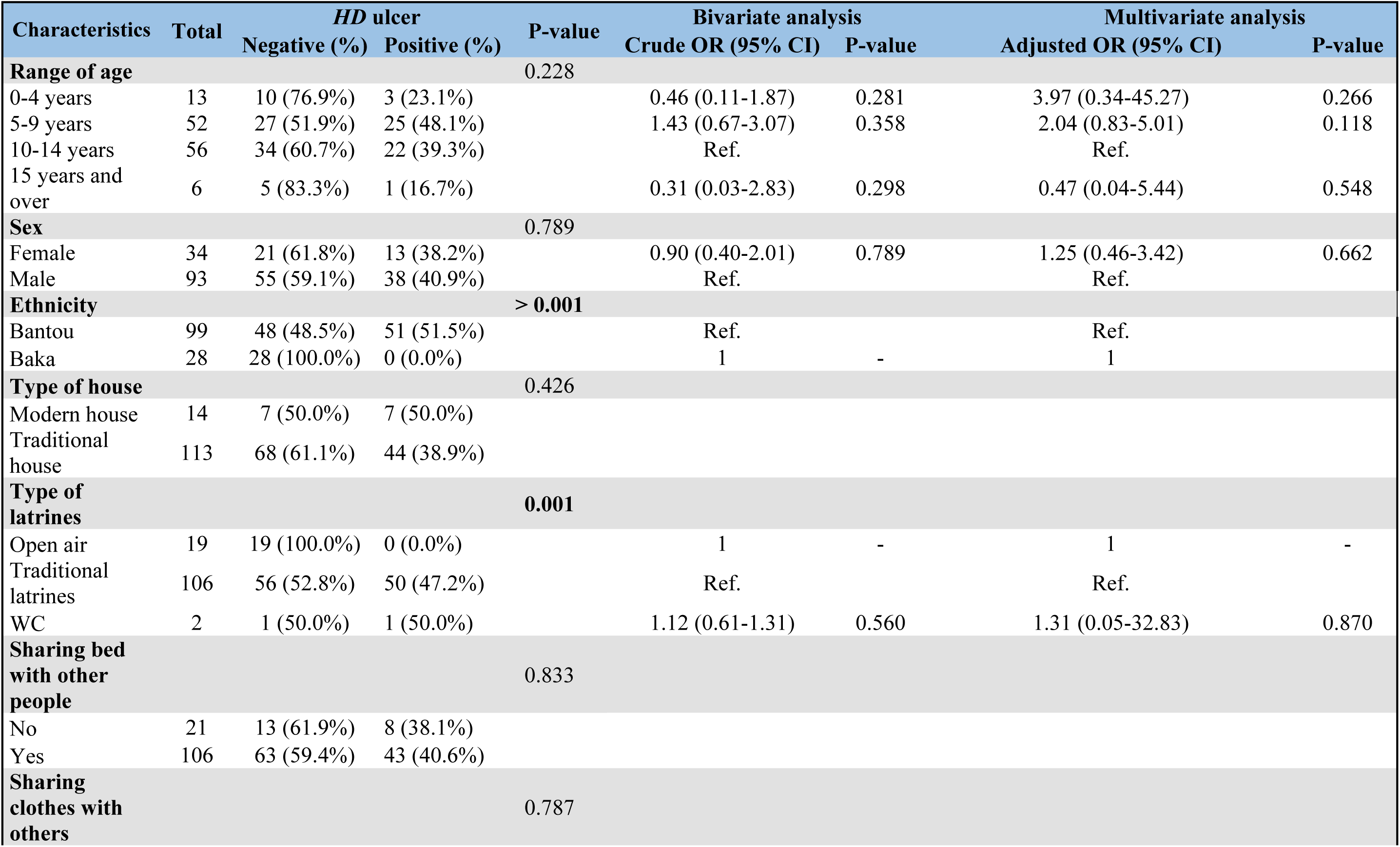
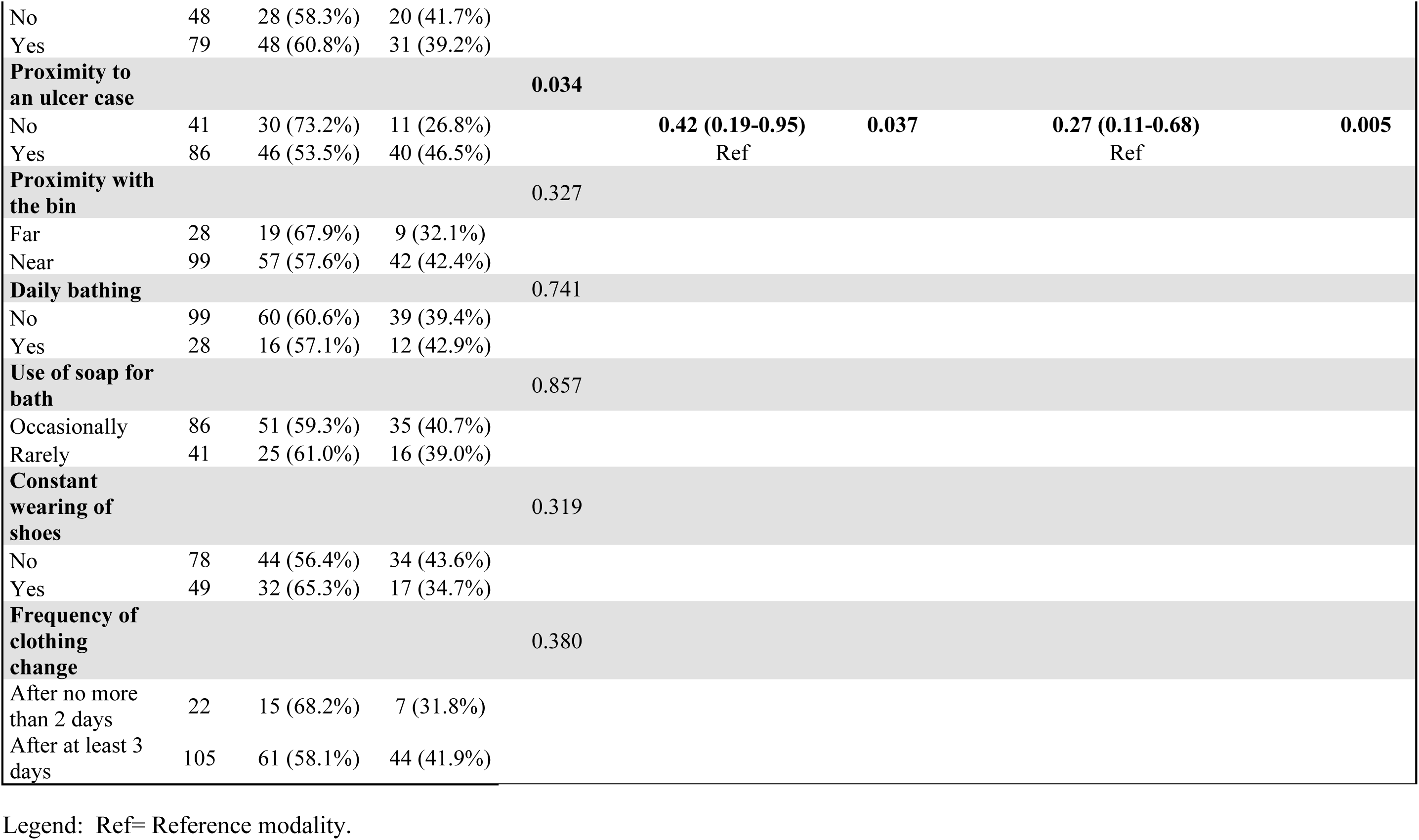
Risk factors associated to *HD* cutaneous ulcers.

**Table 4.**
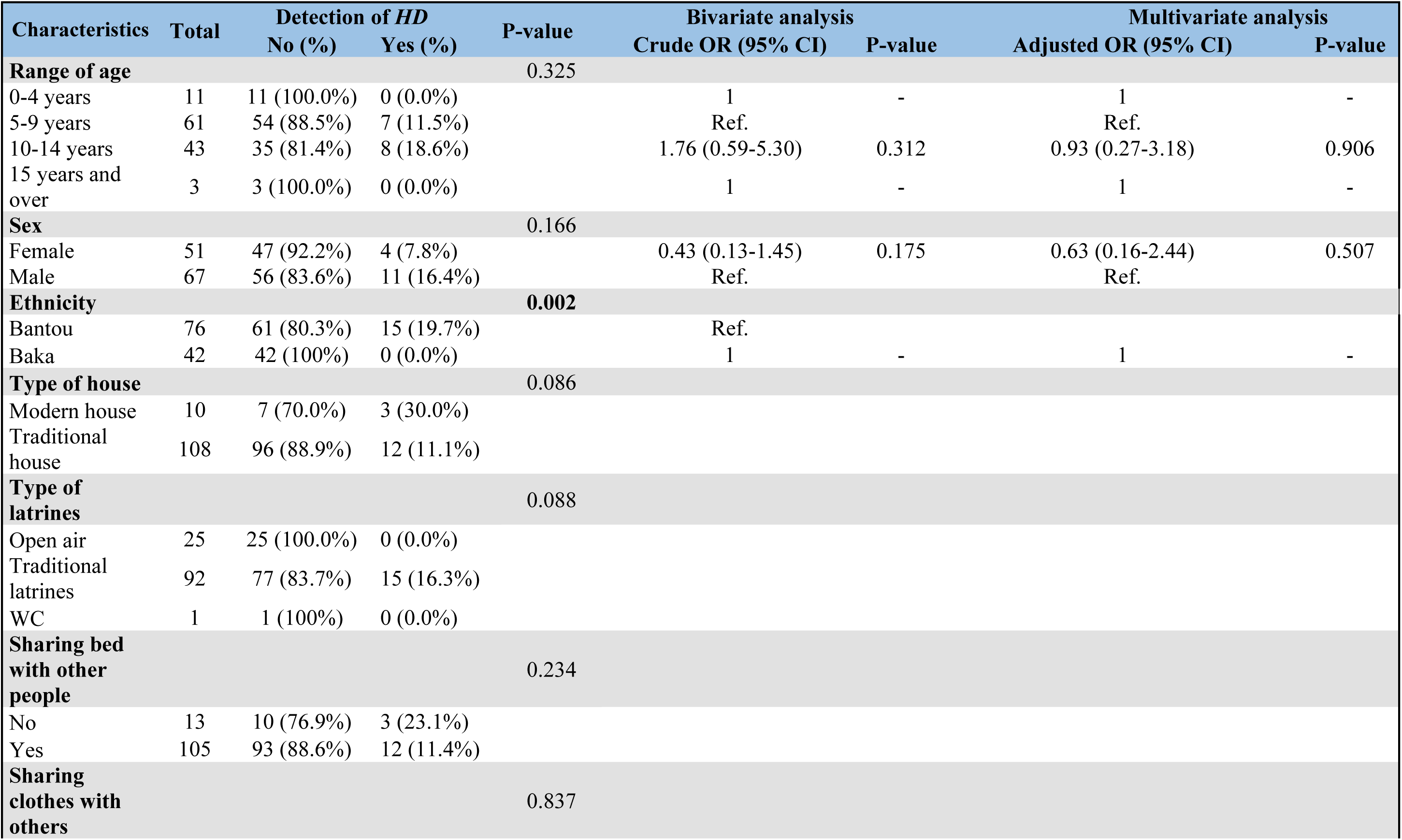
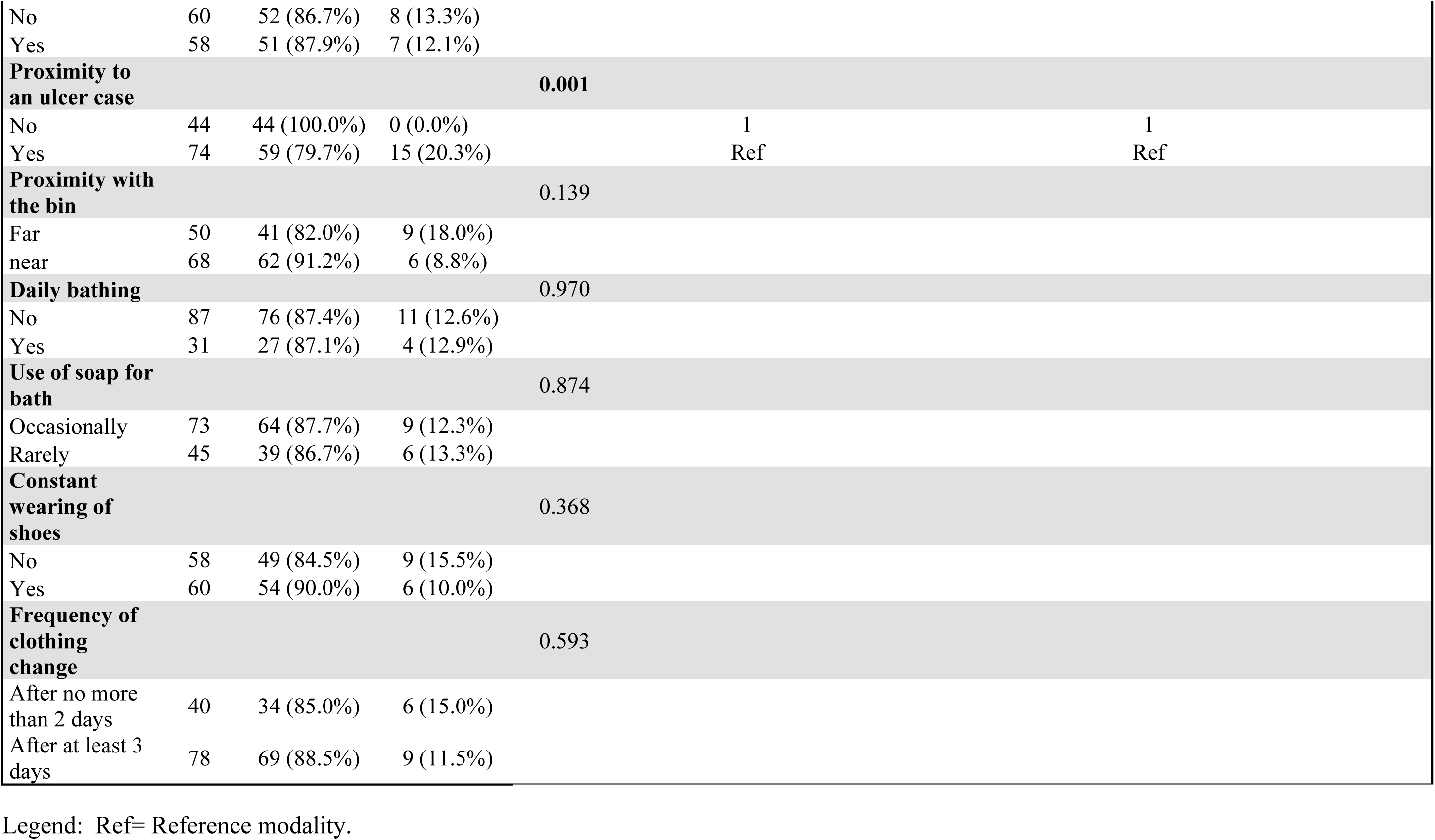
Risk factors associated to the carriage of *HD*.

## Discussion

In this comprehensive study we found that approximately 1% of the screened population in Cameroon had a cutaneous ulcer and that over 30% of these were associated with *HD* infection, making it the single commonest causative organism identified. Ulcers were more frequent in children and appeared to be related to several parameters regarding sanitation and hygiene. We also detected asymptomatic colonisation with *HD* and this occurred exclusively in people exposed to an individual with *HD* ulcer.

Consistent with some previous studies we found the proportion of cutaneous ulcers associated with *HD* (30.3%) was much higher than those associated with *TPE* (5.2%). Our previous surveillance-based study in Cameroon showed that *HD* was responsible for almost half (49.6%) of yaws-like ulcers identified between 2017 and 2019, slightly more than twice those caused by *TPE* [17]. In this study, some of the samples were collected from participants who had washed and disinfected their ulcers prior to collection, which could have led to degradation or low load of *HD* DNA. The previous study also predominantly involved investigating reported outbreaks of cutaneous ulcers whilst the current study was conducted in normal conditions in the community which may also have impacted on our findings.

We identified a number of sociodemographic characteristics alongside markers of poor personal hygiene and sanitation behaviours that were associated with an increased risk of cutaneous ulcers. Apart from a study in 2017 that did not find a significant association between age and cutaneous ulcers [29], our results are consistent with the findings of a case-control study in Ghana where raising age was associated with increased odds of infection [25]. The practice of activities likely to induce trauma or scratches such as farming, animal husbandry and rough play tends to increase with age [30]; these smaller wounds could later serve as a gateway to the bacteria responsible for infection [31]. We found the proportion of men affected by cutaneous ulcers was slightly higher than that of women which has been reported elsewhere. This may be partially attributed to host biological factors [32] and to a higher risk of microtrauma amongst boys and men. Ethnicity was the only socio-demographic variable studied associated both for general cutaneous ulcers and for *HD* ulcers, with Bantu people being more likely to get infected by *HD* cutaneous ulcers compared to Baka people. These unexpected results contrast with several previously published reports. Most of the cutaneous ulcers which correspond to bacterial ulcerative conditions affect preferentially poor people living in warm, moist climates and mainly in forested tropical regions [33,34]. This correspond to the natural environment of the Baka population of Central Africa which until then constituted the most at risks community in this region [35–38].

We found people in close and constant contact with other cutaneous ulcer cases were three times more likely to be infected and most of the cases of *HD* ulcers were detected in schools, among classmates. This school environment is characterised by close proximity and frequent contact between children which could favour the transmission of *HD.* Nearly all individuals with an *HD* ulcer made use of traditional latrines characterized by cracked mud walls, damp earthen floors and lack of roof. These kinds of toilets might facilitate interactions with flies which had been identified as possible mechanical vector for the transmission of *TPE* and *HD*. DNA of *HD* has been detected in flies collected from areas immediately outside the houses of individuals with cutaneous ulcers [21]. We also found people who shared their clothes with others or only changed clothes infrequently had an increased risk of cutaneous ulcers which is consistent with previous studies showing a higher risk among people who share personal items such as towels, clothes and sponges [25,39,40].

*HD* was detected in 15 (8.7%) individuals without cutaneous ulcers which is about half the rate seen in a previous study [21]. In the current study all asymptomatic individuals in whom *HD* DNA was detected were linked to a confirmed case of *HD* cutaneous ulcer whereas in the previous study *HD* DNA was found in almost the same proportion in asymptomatic individuals exposed and not exposed to ulcers [21]. *HD* spreads naturally through skin contact [33,41], and so far no real evidence of an alternative transmission route has been clearly demonstrated. In this context, it seems plausible that the *HD* DNA detected in asymptomatic individuals results from direct or indirect contact with confirmed ulcer cases.

Our study had some limitations. Firstly, because of the COVID-19 pandemic, we could not visit the entire population of the community with many schools and households declining to undertake the preliminary screening step of inclusion and this could have affected the representativeness of our study population. We relied predominantly on detection of DNA and not culture, which is technically challenging, and our data are cross-sectional in nature. As a result, further studies are needed to draw more robust conclusions about the role of asymptomatic colonization.

Our data confirm that *HD* is a leading cause of cutaneous ulcers in yaws endemic health district in Cameroon and is associated with limited access to water and sanitation. Whilst we confirmed the evidence of *HD* colonization have further studies on its viability and implications for transmission should be undertaken to help inform control strategies.

## Data Availability

All relevant data are within the manuscript and its supporting information file

## Acknowledgements

This study is part of the Lamp4Yaws EDCTP funded project (grant number RIA2018D-2495 (grant number RIA2018D-2495). We would like to thank the field team of this study. We thank all the study participants and their legal representatives for their time.

## Supporting information

S1 file. Distribution of *H. ducreyi* and *T. pallidum* positive participants per site. Legend: CU= cutaneous ulcer case; AS= asymptomatic individual

